# The spatio-temporal distribution of musculoskeletal disorders: results of the Global Burden of Disease in 204 countries and 21 subregions between 1990 and 2019

**DOI:** 10.1101/2022.09.16.22280040

**Authors:** Hanifa Bouziri, Yves Roquelaur, Alexis Descatha, William Dab, Kévin Jean

## Abstract

**Objectives:** This study aimed to globally assess the prevalence and distribution of primary-origin musculoskeletal disorders (MSDs) from 1990 to 2019 to better understand their temporal trends.

**Methods:** Using data from the 2019 Global Burden of Diseases, prevalence rates of 6 primary-origin MSDs were analysed across sub-regions, age groups, and genders. Raw and age-standardized data were mapped for over 204 countries. Cochran-Armitage trend tests evaluated temporal prevalence trends. The correlation between MSDs prevalence, national income levels, and medical density was explored.

**Results:** In 2019, global MSDs prevalence varied significantly among countries. Hip osteoarthritis had a prevalence of 0.56% [95% CI: 0.43-0.70], while low back pain was 8.62% [95% CI: 7.62-9.74]. Most MSDs exhibited an increasing prevalence with age, except for neck pain, which stabilized or decreased after age 45-50. Women generally had higher prevalence rates across all age groups. High-income countries consistently showed higher prevalence rates compared to middle and low-income countries. Over time, most sub-regions experienced a significant increase in MSD prevalence. However, after adjusting for age, the temporal trends for back and neck pain became non-significant, except for hip osteoarthritis, where half of the sub-regions remained significant. Multivariate linear regressions revealed positive associations between MSD prevalence and both national income level and medical density.

**Conclusion:** The global burden of MSDs is increasing due to population ageing, but other factors should be considered. Longitudinal studies with a wider range of MSDs and additional risk factors are needed for improved prevention strategies.

## Introduction

Musculoskeletal disorders (MSD) are among the most common conditions worldwide, affecting 1.71 billion people in 2020. MSD includes approximately 150 distinct conditions and can affect people’s quality of life, healthcare costs, and work efficiency [1]. Beyond health issues, MSD may generate considerable productivity loss. In 2010, it was estimated that 39.2% of European workers suffered from chronic pain and could not work to their total capacity [2]. Despite the widespread perception of a Northern country-specific issue, MSD represents a global health concern. In 2016, the Global Burden of Disease (GBD) estimated that low back pain (LBP) was the leading cause of years lived with a disability in 160 countries [3,4]. Providing a detailed picture of the current global burden of MSD and anticipating their likely future trends is thus crucial to assessing the needs of prevention and care and health expenses. However, evidence of past and current trends in the occurrence of MSD comes primarily from high-income countries, while evidence in the Global South still needs to be made available [5].

Several factors may drive the spatial and temporal distribution of MSD. First, several risk factors for MSD have been consistently identified, including occupational exposures such as biomechanical or psychosocial factors [6,7]. Moreover, estimates of MSD burden are likely affected by under-report, which may be driven by diagnostic capacities and cultural factors, such as perceptions of MSD by caregivers and patients [6,8]. Therefore, trends in these individual, occupational, or diagnostic-related factors may contribute to the temporal and spatial distribution of MSD. Age also constitutes a strong risk factor for MSD. Among the working population, the MSD incidence is globally higher in people over 50 years [9,10]. Thus, the global variations in demographic structures may also affect the spatial distribution of MSD. Similarly, the global demographic trend of ageing populations may also impact the temporal trends in the burden of MSD [11,12]. Assessing the contribution of demography in MSD’s past trends may thus be insightful to anticipate their fate regarding projected further population ageing.

A global view of the geographic and temporal trends in MSD occurrence still needs to be improved. This study aims to understand the spatio-temporal distribution of MSD worldwide, between 1990 and 2019, for 204 countries and 21 territories, based on an analysis of the GBD 2019 study.

## Method

### Data sources

The GBD is the largest and most comprehensive effort to measure global disease prevalence levels and trends over time [13]. The GBD is coordinated by the Institute for Health Metrics and Evaluation (IHME). The collection of health data as well as the characteristics of individuals, is based on medical claims, general population surveys and medical diagnoses [14]. In addition to collecting data on health events by age, sex, and geographical area over time, the GBD produces regular point estimates of the indicators studied with their 95% uncertainty intervals. The detail of the GBD protocol is accessible on their website [15]. For our study, we accessed 1990 to 2019 GBD data at: https://vizhub.healthdata.org/gbd-results/ [16]. Methods for estimating all the accessible morbidity have been described more fully elsewhere [17,18]. In brief, all available global data, including vital registration, sample registration, household surveys, censuses, disease registries, reporting systems and police records were identified, extracted, and normalized. Standardized methods were then applied to produce consistent estimates of the general population of each country considering fertility, net migration, all-cause mortality, and cause-specific mortality.

National income levels were measured using the World Bank income category provided by the GBD, analysed as an ordinal variable (high-/upper-middle-/lower-middle-/low-income country) [16]. The national 2017 values of medical density are expressed as the number of healthcare providers per 1,000 inhabitants extracted from the World Bank’s BIRD - IDA database [19].

### Case definition

The present study focuses on MSD of primary origins, for which it is possible to consider primary or secondary prevention policies. Therefore, we excluded MSD from inflammatory origin or resulting from a joint manifestation of organic diseases (such as gout, lupus, psoriasis, certain infectious diseases, etc.). We thus included the following definitions, based on two types of MSD data collection: self-reported pain and medically diagnosed osteoarthritis. Both were used for distinct sites: i) low-back pain, ii) neck pain, iii) hip osteoarthritis, iv) knee osteoarthritis, v) hand osteoarthritis, and vi) other osteoarthritis.

Low back pain (LBP), neck pain (NP), and osteoarthritis (OA) were classified according to the International Classification of Diseases (ICD-10). The “other osteoarthritis” category represents the most common form of arthritis, involving chronic inflammation, breakdown, and structural alteration of the joint. Here, the reference case was radiographically confirmed, symptomatic OA in any joint other than the hand, hip and knee treated independently [17–19].

Data collection covered 204 countries and 21 sub-regions between 1990 and 2019 [16]. These subregions were further classified into six larger regions: the Americas, the Caribbean, Europe, Oceania, Asia, and Africa. We selected age groups between 20 and 70 years old.

### Analysis

#### Spatial and temporal distribution of the prevalence of MSD across countries and territories, 1990-2019

For each MSD studied, we mapped national raw and age-standardized prevalence (supplementary materials S1). Then we analysed temporal trends of raw and age-standardized prevalence of MSD, by sub-regions and globally, by computing annual population-weighted average prevalence with the corresponding 95% confidence interval (95% CI). Temporal trends in raw and age-standardized prevalence were tested using Cochran-Armitage (CA) trend tests. Analyses were conducted on males and females first and then stratified by sex.

#### The distribution of the prevalence of MSD considering the income level and medical density worldwide

We hypothesised that country-level prevalence of MSD was correlated with national income level, which may can be considered as a proxy capturing rough inequalities between countries and indirectly reflecting the level of economic development and relative wealth of a country. Additionally, for medical density we considered that this indicator may capture MSD diagnostic capacities.

This variable was categorised based on the quartiles of its distribution. The independent effect of income level and medical density was explored using multivariate linear regression. All analyses were conducted using the R software (v 4.1.1).

## Results

### Spatial and temporal distribution of the prevalence of MSD by worldwide countries and territories between 1990 and 2019

In 2019, the global raw prevalence mean of MSD were ranged from 0.56% [95% CI: 0.43-0.70] for hip osteoarthritis to 8.62% [95% CI: 7.62-9.74] for low back pain, with significant variations across countries (S2). More specifically 0.46% [95% confidence interval, CI: 0.35-0.58] for hip OA, 1.71% [95% CI: 1.27-2.29] for hand OA, 2.34% [95% CI: 1.84-2.98] for neck pain, 4.34% [95% CI: 3.71-5.02] for knee OA and 8.22% [95% CI: 7.21-9.35] for low back pain. Country-specific prevalence ranged from 0.04% [95% CI: 0.02-0.05] for hand OA in Timor-Leste to 18.9% [95% CI: 16.7-21.4] for LBP in Japan, with important variations across MSD and regions (**S3**). Among the 6 MSD analysed, lower prevalence levels were consistently observed for sub-Saharan Africa.

The higher overall average prevalence (%) per sub-regions all year combined was observed in **Figure 1**. They varied between 0.09% [95% CI: 0.06-0.13] for hand OA in Southeast Asia and 15.8% [95% CI: 14.6-17.3] for low back in High-income North America. The lowest overall prevalences (%) per sub-regions all year combined were for the LBP in Eastern Sub-Saharan Africa at 3.9% [95% CI: 3.4-4.4], for NP in Central Sub-Saharan Africa at 0.7% [95% CI: 0.5-0.9], for hip OA in Eastern Sub-Saharan Africa at 0.1% [95% CI: 0.09-0.15], knee OA in Eastern Sub-Saharan Africa at 1.4% [95% CI: 1.1-1.6] and the hand OA in Southeast Asia at 0.09% [95% CI: 0.06-0.13]. The highest overall prevalences (%) per sub-regions all year combined were for NP in Western Europe at 4.7% [95% CI: 3.8-5.8], hip OA in Western Europe at 1.2% [95% CI: 0.9-1.6], knee OA in High-income Asia Pacific at 9.7% [95% CI: 8.5-11.0] and for the hand OA in Eastern Europe at 7.2% [95% CI: 5.5-9.4]. The overall average prevalence for all years combined has been observed to be higher in high-income countries for LBP, hip, knee, and other OA. For overall prevalence, in all years combined, NP and hand OA were more elevated in high- and middle-income countries (**Figure 1**).

**Figure 1.**
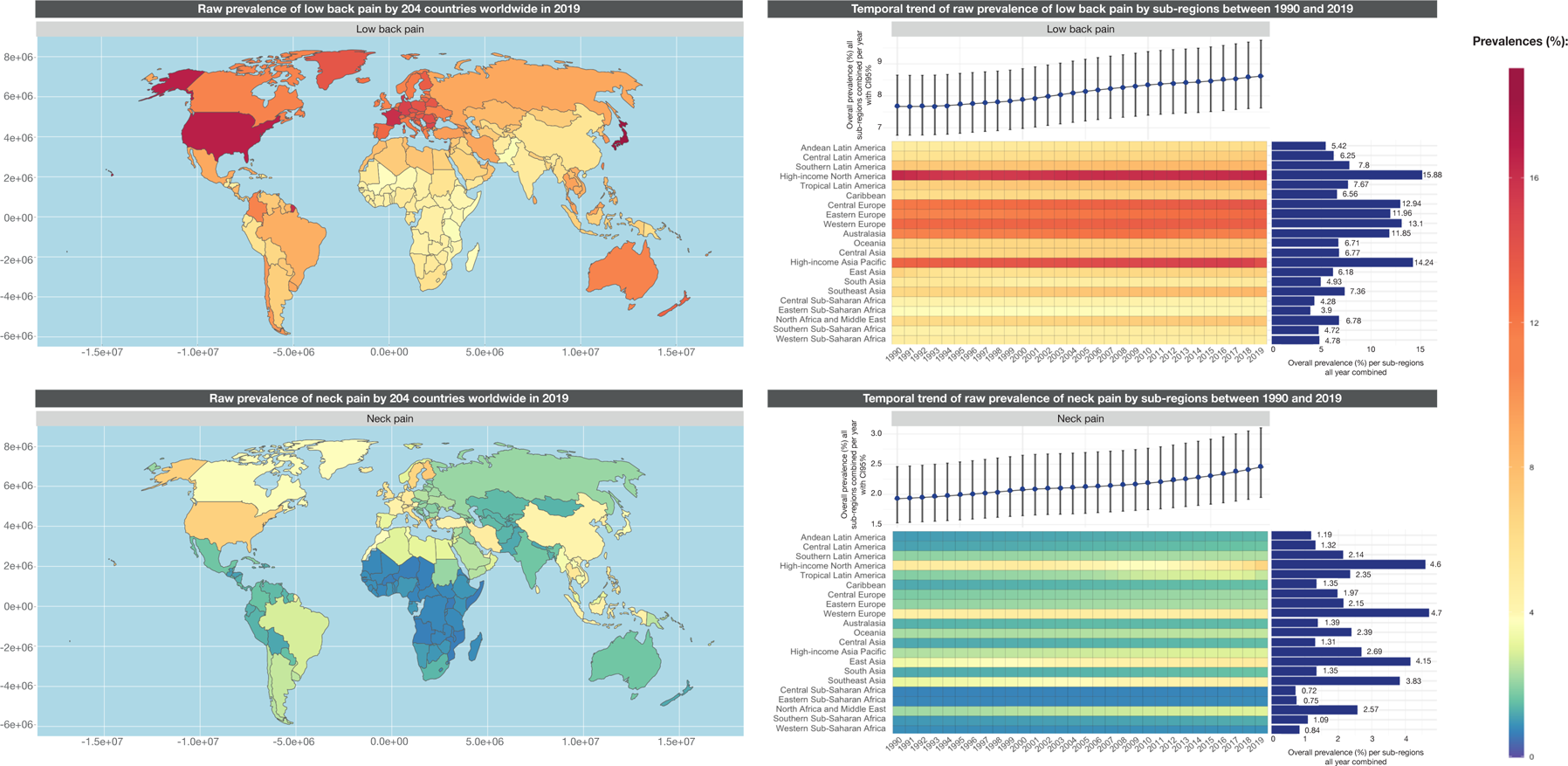
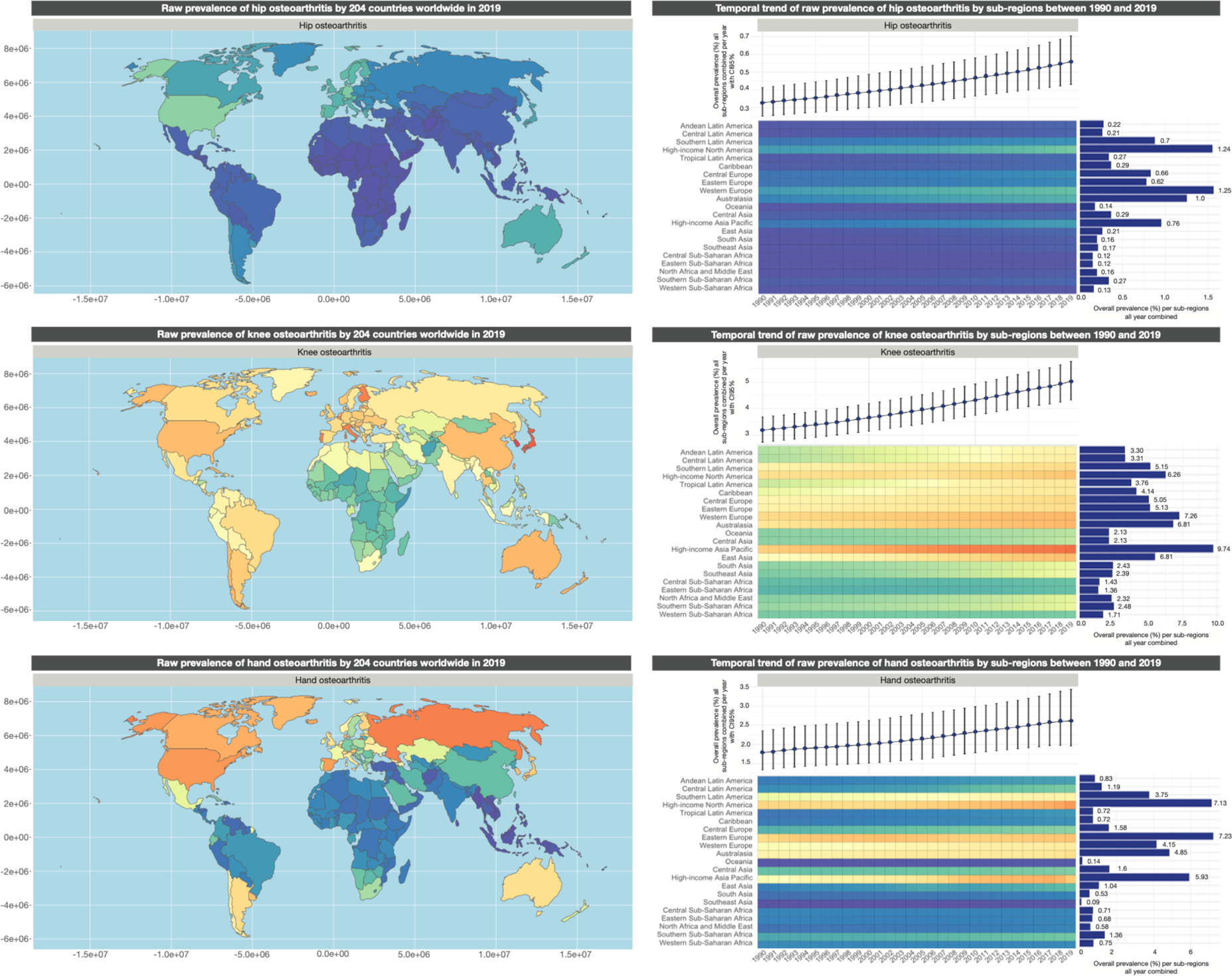
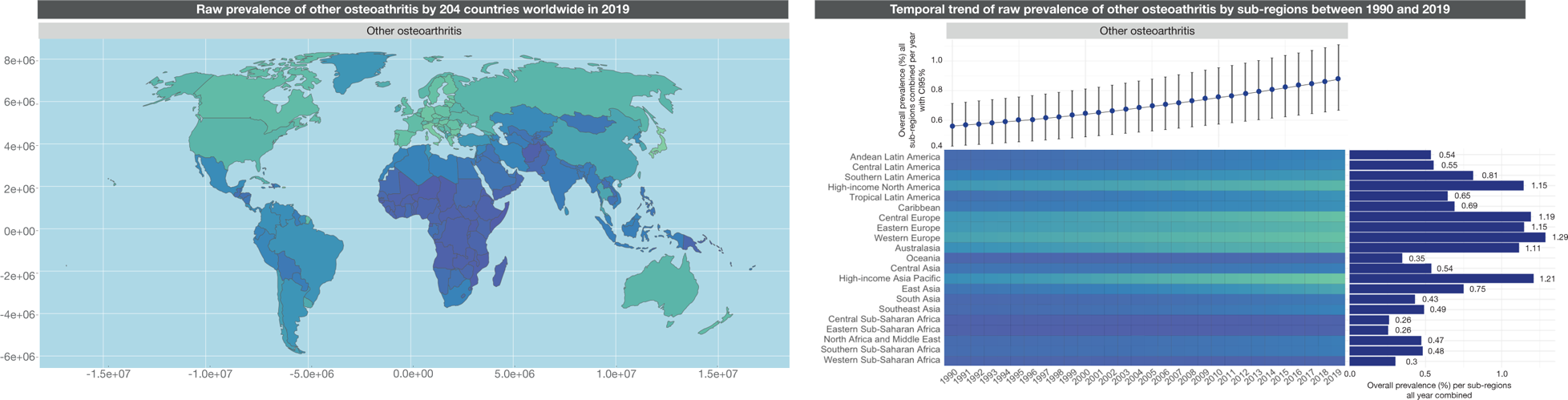
Spatial and temporal distribution of the raw prevalence of MSD worldwide by 204 countries and 21 sub-regions between 1990 and 2019.

Globally, the prevalences of LBP, hip/hand OA and other OA increase with age in both males and females (**Figure 2**). The 2019 age-specific prevalence of neck and knee OA increased, then peaked around age 45 for NP and 55 for knee OA. Although absolute levels varied, the shapes of the sex-specific age distributions were similar across sub-regions. Prevalences were higher in women, except for hip OA in Southern Sub-Saharan Africa and other OA, which prevalence was higher in men.

**Figure 2:**
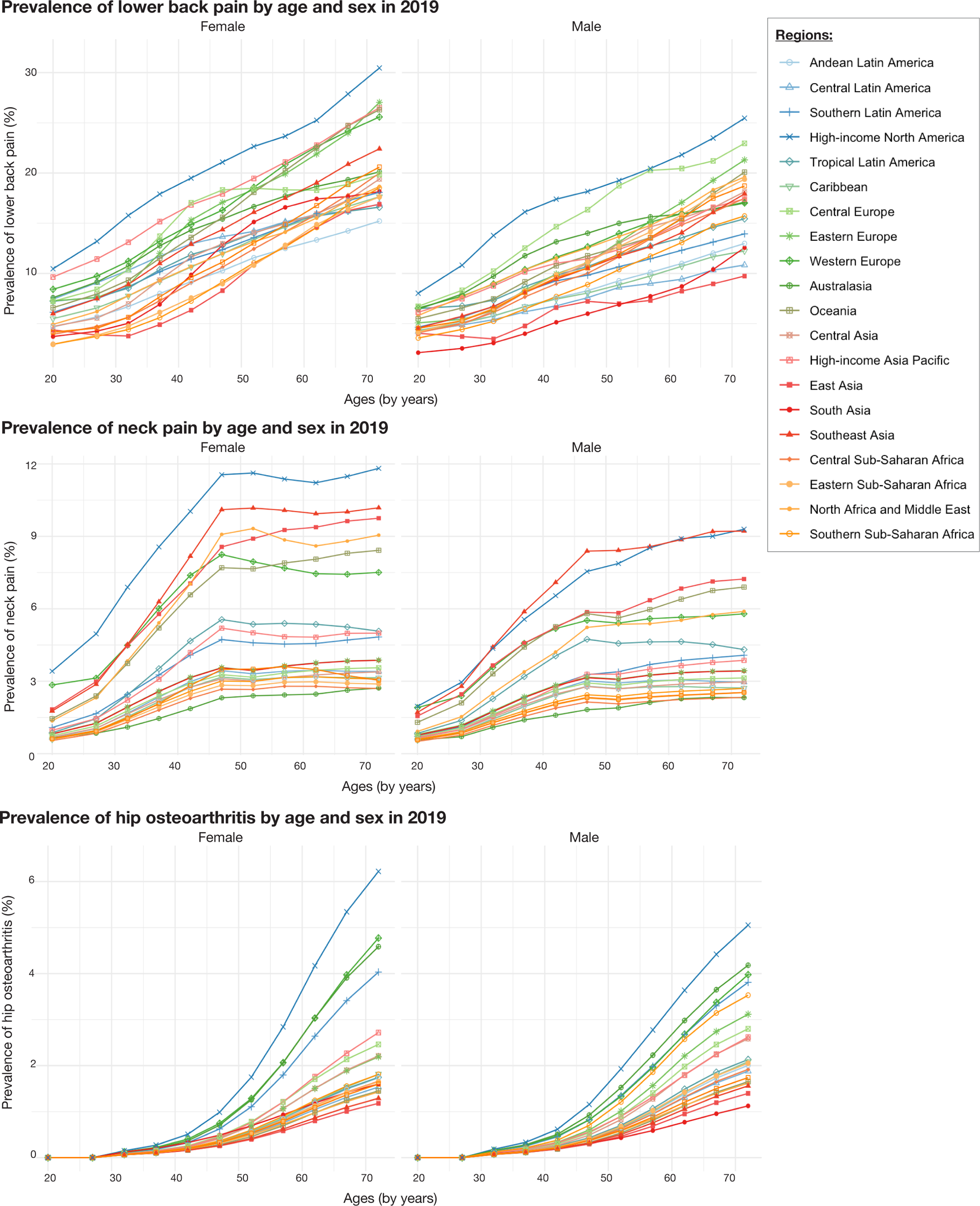
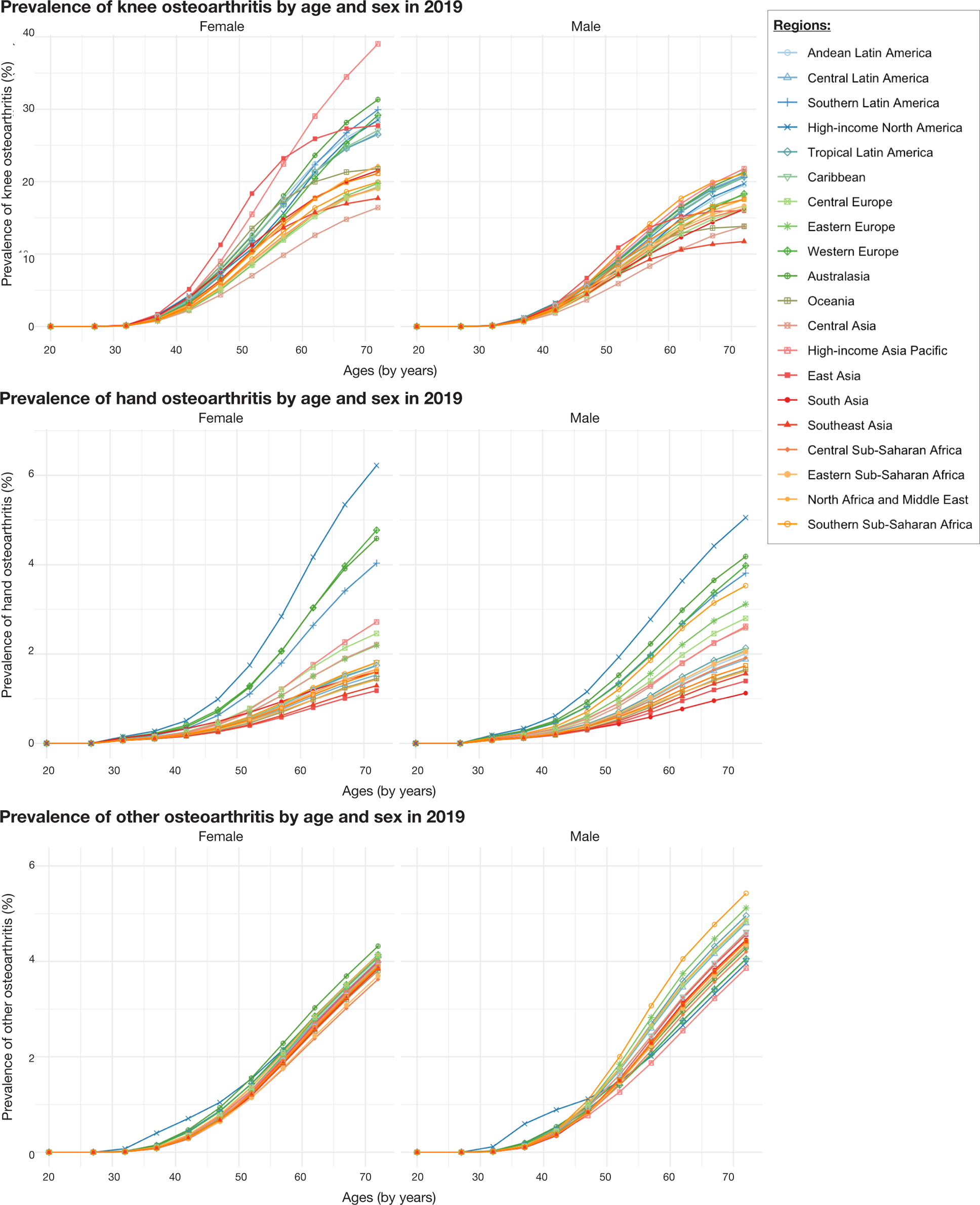
Prevalence of MSD worldwide by sub-regions, age, and sex for 2019.

Over time, more than half of the raw prevalence of pain increased significantly. In contrast, after age standardisation, the trends were mainly non-significant except for four decreasing pain trends: 3 for LBP in Australasia, East Asia, and South Asia and 1 for NP in high-income North America (**Table 1**). As observed for LBP and NP, most osteoarthritis was explained by hand and other OA demographics. However, unlike LBP and NP, nearly half of the increasing trends remained so after age-standardization for hip OA (13/22 significant or near-significant increase) and knee OA (6/22 significant increase or near-significant increase).

**Table 1:**
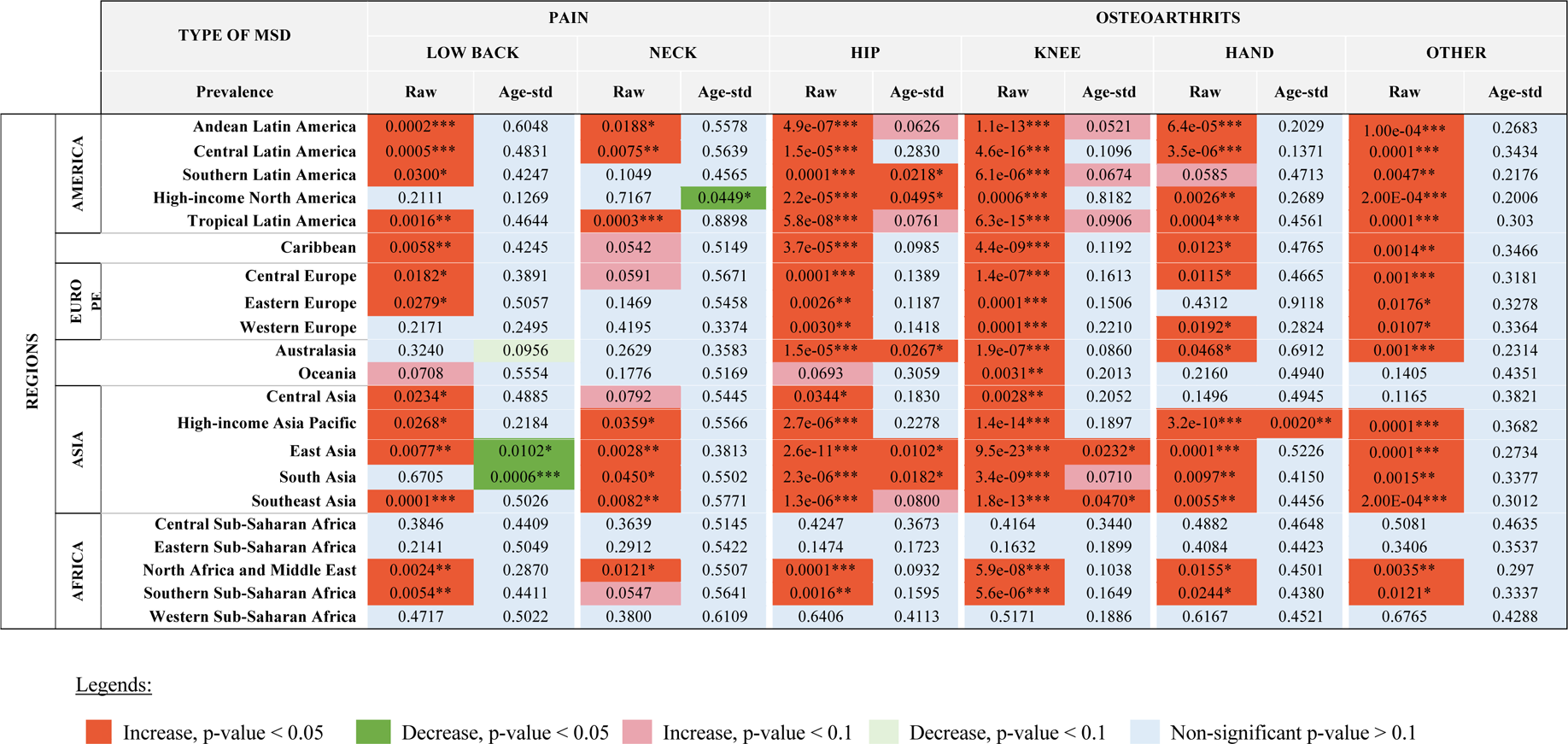
Cochran-Armitage trend tests on the raw and age-standardized prevalence of MSD between 1990 and 2019, by regions and sub-regions in both sexes.

To compare temporal trends in MSD prevalence by sex, 252 CA trend tests were performed for age-standardized prevalence. Among these tests, differences were observed (**S4**) for 21 sub-regions. For LBP, three different temporal trends were highlighted when considering age: in Australasia, the trends are stable for women and decrease significantly in men; in East Asia and South Asia, trends increase significantly for women and reduce significantly for men. There are no differences in time trends of age-standardized MSD for NP. For hip OA, eight temporal trends differences were observed between women and men: in Andean Latin America, Southern Latin America, High-income North America, Tropical Latin America, Australasia, and Southeast Asia, stable trends for women and increasing trends for men were highlighted, while for East Asia and South Asia, the decrease was observed in women and an increase was observed in men. For knee OA, five temporal trends in the prevalence of MSD considering age were noticed: in Central Latin America, Tropical Latin America, Caribbean, and Southern Sub-Saharan Africa, the trends were stable for women and increased in men, while in South Asia, the opposite was seen. Finally, for hand OA, two differences in trends were observed: in Eastern Europe, the trends were stable in women and decreased in men, while for High-income Asia Pacific, in women, the trends increased while in men were stable.

### The distribution of the prevalence of MSD considering the World Bank incomes and the medical density worldwide

We observed a positive association between the prevalence of MSD for the World Bank income level and medical density (**Figure 3**). In multivariate regressions, both variables were independently positively associated with the prevalence for each of the 6 MSD (**S5**).

**Figure 3:**
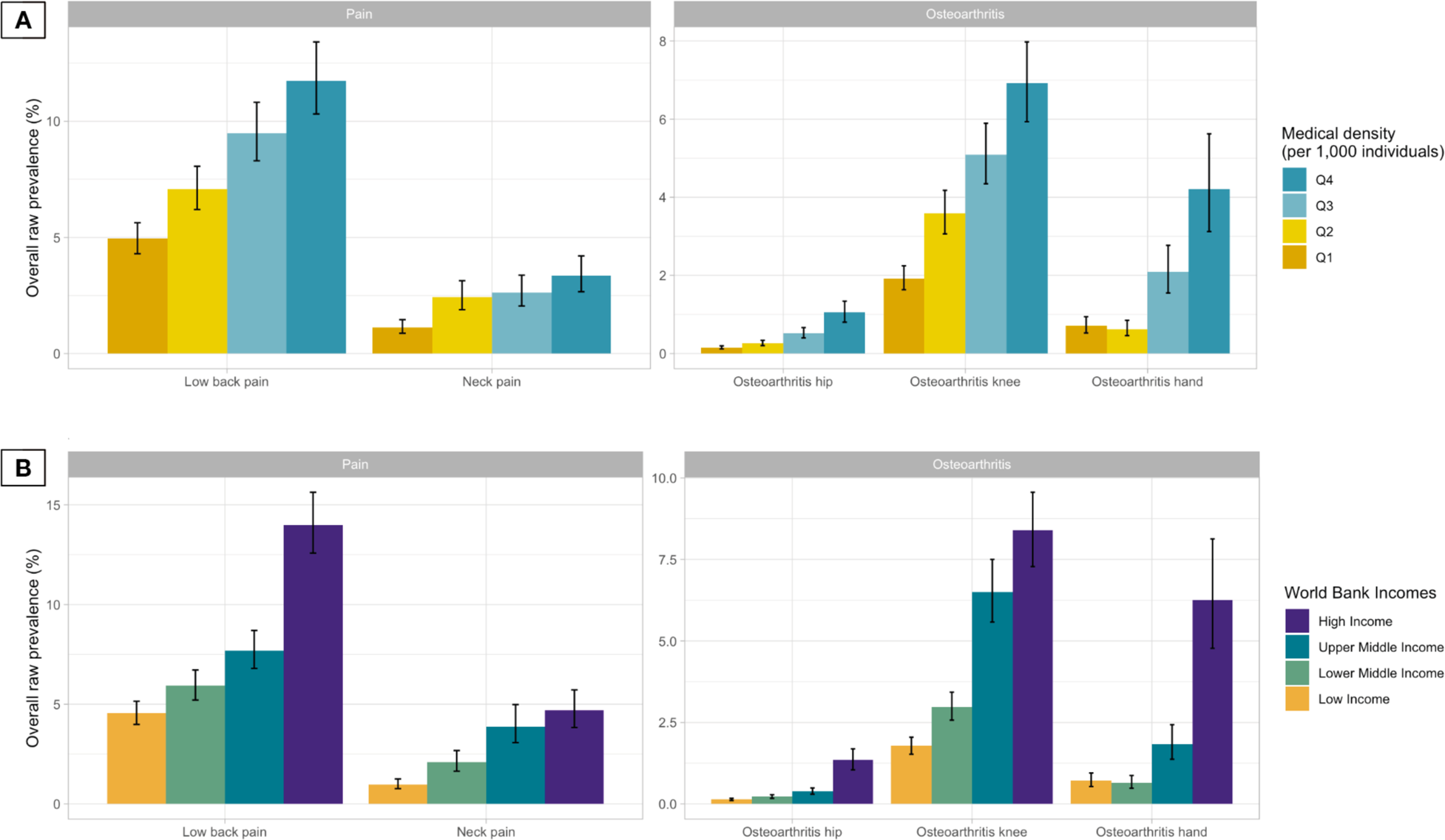
The distribution of the prevalence of MSD considering the World Bank incomes and the medical density worldwide. **A.** Distribution of MSD prevalence by World Bank income (low, lower-middle, upper-middle, and high incomes) in 2019. **B.** Distribution of MSD prevalence by medical density (per 1,000 individuals) categorised by quantiles in 2017.

## Discussion

In this study, relying on an extensive analysis of 6 MSD of primary origins, we describe several patterns driving their spatial and temporal distribution. First, we observed large geographical variations in the raw prevalence of MSD across sub-regions, with a constant trend toward higher prevalence in high-income countries. Second, we observed that the prevalence of MSD increases monotonically with age, except for neck pain, which plateaus or decreases after 45-50 years old. Third, we report that for 5 out of the 6 MSD we studied, women were more affected than men, constantly across sub-regions. Fourth, we document that globally, the raw prevalence of MSD has significantly increased between 1990 and 2019, with some variations across sub-regions. This increasing trend is likely to be mainly driven by the ageing of the population, as age-standardized prevalence remained relatively the same over time. Fifth, when studying the drivers of the spatial distribution of MSD, we observed that large variations remain in prevalence levels when controlling for age, suggesting that demographics only cannot explain spatial disparities in MSD prevalence. Sub-regional levels of income level and medical density were independently associated with a higher prevalence of MSD, suggesting that both labour landscape and diagnostic capacities may affect the local prevalence levels. Furthermore, it is also interesting to note that a probable under-reporting of MSD can partly explain the low prevalence values in low-income countries since, in these countries, research priorities are often directed towards pathologies or health problems requiring rapid action (undernutrition, issues related to geopolitical conflicts, infectious diseases, work-related death or another competitive risk etc.), whereas high income is more on long action (overnutrition, psychosocial …).

A significant disparity in overall average prevalence between the sub-regions, all years combined, depending on the MSD, has been observed. For LBP, the difference is a factor of 4, with a 3.9% prevalence in Eastern Sub-Saharan Africa and a 15.8% prevalence in high-income North America. For NP, the prevalence in Western Europe (4.7%) is more than six times higher than in Central Sub-Saharan Africa (0.72%). For hip OA, in Central and Eastern Sub-Saharan, the prevalence is 0.12% which is ten times lower than for Western Europe, which has a prevalence of 1.2%. The range between the highest and lowest prevalence for knee OA is more than 7, with a prevalence of 1.3% for Eastern Sub-Saharan Africa and a prevalence of 9.7% for the High-income Asia Pacific. Finally, the prevalence of High-income Asia Pacific at 7.2% is about 80 times higher than for Southeast Asia, which has an overall average prevalence for all years combined of 0.09%. Differences in demography do not solely explain these significant variations in prevalence, as large prevalence differences remain while considering age-adjusted prevalence.

This study showed that the most widely high prevalence is attributable to LBP, corroborating previous results from a study published in 2014 [22]. The raw prevalence of MSD increased between 1990 and 2019 in most sub-regions of the world, but most of the pains were explained after considering the age factor, while there were still persistent increases for hip, knee, and other OA. Depending on the type of MSD, demographic seems to explain most temporal changes in pain. Still, more is needed to understand and apprehend its geographic distribution correctly. Another interesting point is that when observing the prevalence of MSD for 2019 by sub-region, sex, and age, most of the prevalence increased with age and at a higher level for women. This higher prevalence in women may be due to several reasons: differential exposures; interactions between exposures and gender; effect modification due to male/ female social roles, genetics, psychology and physiology; differential pain experience, reporting or care-seeking [23]. These results improved our understanding of temporal trends in MSD, suggesting that the overall average prevalence of MSD will increase globally over time. Furthermore, in regions where comprehensive income and medical density are lower, the burden of MSD was ranked lower, probably due to a lack of means and accessibility to diagnosis. Added to this is that in recent years, the world population has been ageing, which implies that a considerable number of people living with MSD is expected in the decades to come [24].

Significant disparities in prevalence between countries have been highlighted, especially a marked distinction between the South and the North countries, with higher prevalence in the latter. After age standardisation, OA increase can be explained by the potential underdiagnosis of patients who do not necessarily report their pain because they consider it sufficiently low. In addition, differences in prevalence can be explained by specific characteristics of the populations, not only by the age pyramids but also by obesity [25], in personal behaviors such as the level of physical activity, the increase of telework [26], and/or in professional exposures such as biomechanical and psychological exposures [6], ergonomics of the workstation and the tools used at work [27], cultural factors in the diagnosis of patients [28], or even in pain tolerance. These aspects mean that the sensitivity and specificity of the indicators observed undoubtedly vary geographically due to the diversity of the countries’ characteristics and the populations’ profiles. Among these variations, we can count the differences in definitions, in national income, reflecting a scarcity in the diagnosis of patients. Another factor that can influence a difference in detecting patients is in the variations in medical density which may be used as an indicator of diagnostic capability and probability of detection [5]. Moreover, the country’s treatment arsenal may also indirectly drive the country’s diagnostic capacity. Another point to consider is the potential improvement in the sensitivity of diagnostic tools and knowledge of these pathologies in North countries. Given these results, the low prevalence in the middle- or low-income countries could see their values increase considerably, given the rise in income per country over time and potentially in medical density and accessibility to care.

The variations in the occurrence of MSD depending on the country can also be explained by the evolution of habits over time, such as whether to practice a physical activity or using transport to get to work. The evolution of professions, the emergence of new jobs, or the differences in the distribution of profiles of individuals by job according to their gender, age or social categories can also play a considerable role in the temporal trends of MSD [29]. Several examples can be cited, such as construction or transport trades mainly occupied by men vs cashiers, caregivers, and housekeepers primarily occupied by women [30,31]. We must consider several opposing factors that can considerably influence the occurrence of MSD. For example, protective factors such as better knowledge of these pathologies with increasingly effective diagnostic and prevention efforts must be considered as much as the known and emerging risk factors. Among the risk factors that will probably increase, we can cite the evolution and emergence of certain professions requiring increasingly rapid activity, leading to high stress and sometimes non-optimal postural and ergonomic conditions, with different levels of risk factors [30–33]. Finally, even if most MSD do not directly lead to death, health is often considerably degraded. Indeed, these pathologies can play a role in premature mortality by inducing combined effects with other pathologies, such as making their diagnosis more difficult [34,35].

Despite the study’s strength, the GBD data present limitations that must be considered in the interpretations for multiple reasons. First, the GBD estimates rely on various data sources in which quality may differ across countries. Second, for certain countries where the data was incomplete, calibration was used on neighbouring countries (mainly observed for African countries) [5]. Furthermore, this study does not allow us to distinguish whether the differences in prevalence observed according to the MSD and the geographical location of the individuals are due to differences in risk which depend on the characteristics between the regions, such as genetic factors, or whether these differences in prevalence are due to variation in occupational or environmental exposure factors.

This study presents other limitations that could explain an underestimation of the prevalence and the real burden of MSD over time 36]. Case definitions for MSD are not universally standardised and challenging in population studies, making analysis sometimes less precise, especially for different incomes by region [37]. Large-scale population studies on the 150 musculoskeletal conditions are challenging to set up due to the exhaustiveness of the pathologies making the setting up of field studies costly [35]. Throughout a person’s lifetime, there can be a wide variation in the likelihood of a condition, and reporting pain resulting from highly variable musculoskeletal diseases sometimes makes diagnosis difficult, especially in low-income areas with low medical density.

## Conclusion

MSD are among the most frequent diseases and the most impactful on the quality of life of individuals worldwide. Globally, there is a significant variation of prevalence between countries and whether the MSD studied is related to pain or OA. The burden of MSD is mainly increasing worldwide, with a great diversity of prevalence between sub-regions. As the ageing of the global population is rising, this temporal trend is expected to continue [38]. Lower back pain is the most widespread worldwide, regardless of the years and locations studied. Over time, the variations in MSD are overwhelmingly increasing, with most of these increases explained by age, particularly for LBP and NP. Demography does not seem to be the only indicator to consider in these variations since, for example, the differences in prevalence observed by country in 2019 highlighted that for all MSD, prevalences are much higher for the North and South countries. In this sense, it has also been shown that income levels and medical densities by region were significantly correlated with observed prevalence. To reduce the burden of these MSD, which tends to increase considerably, it is essential to continue to set up broad surveillance and refine the collection methods by facilitating access to data on a more comprehensive type of MSD [36]. Exhaustive longitudinal data on a larger panel of MSD and relevant factors, considering risks factors over time, would allow us to understand their evolution better and develop efficient policies for managing and preventing these diseases [35,39].

## Data Availability

All data produced are available online at the global burden of disease site

http://ghdx.healthdata.org/gbd-results-tool

## Acknowledgements

The authors are responsible for the opinions expressed in this article, and they do not necessarily represent the views, decisions, or policies of the institutions with which they are affiliated. The authors declare no competing interests and acknowledge funding from the Cnam-Malakoff Humanis “Health and Companies” Chair. The funder has no role in the study design, data collection and analysis, publication decision, or manuscript preparation.

## Contributor statements

K.J., W.D. and H.B. designed the project. K.J. and W.D. supervised the project. H.B., W.D. and K.J. designed the analysis plan, with inputs from A.D. and Y.R. H.B. extracted and analysed the GBD data. H.B. wrote the first draft of the manuscript. All authors revised and commented on the manuscript and approved the final version.

## Supplementary materials

**S1:**
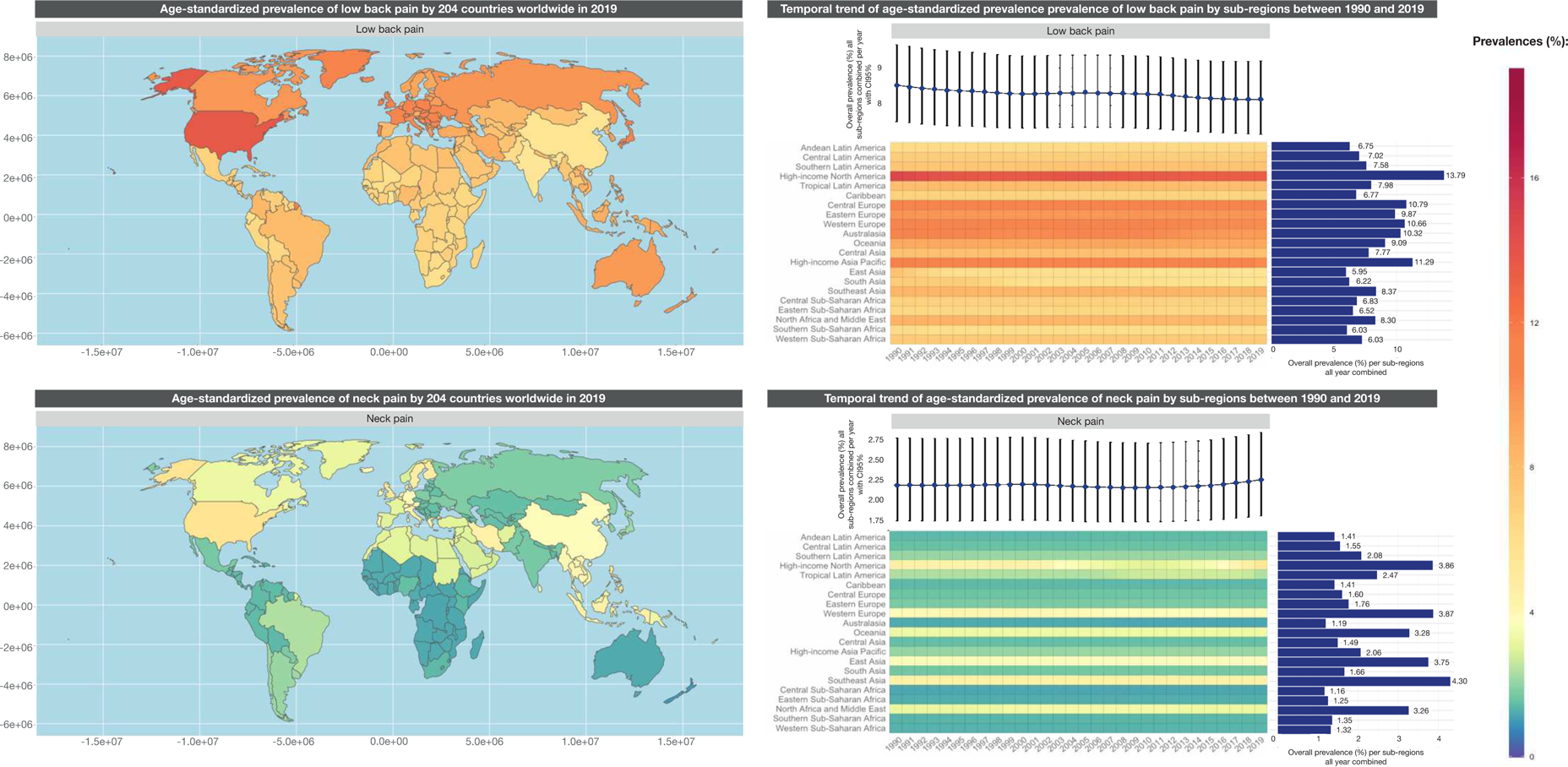
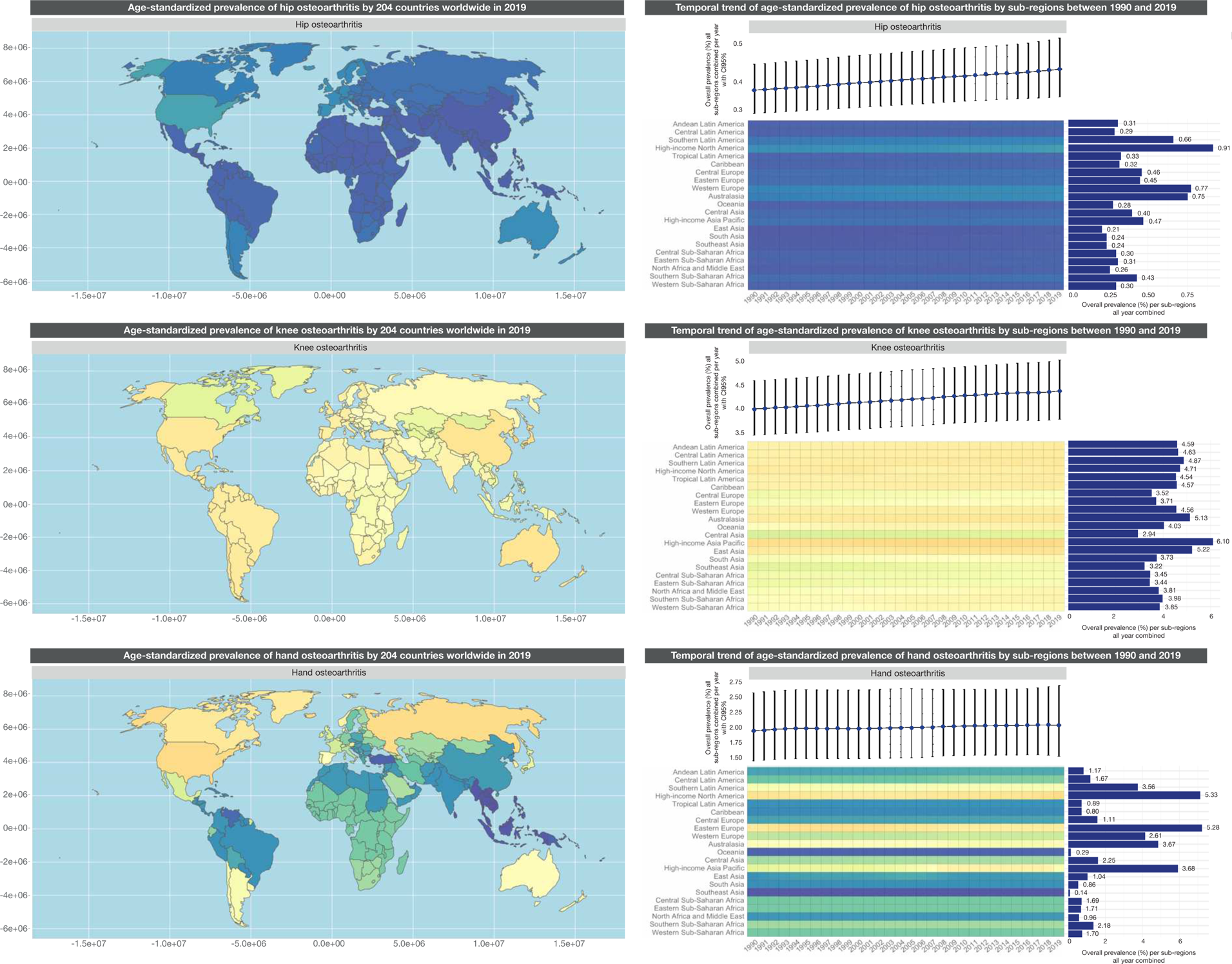
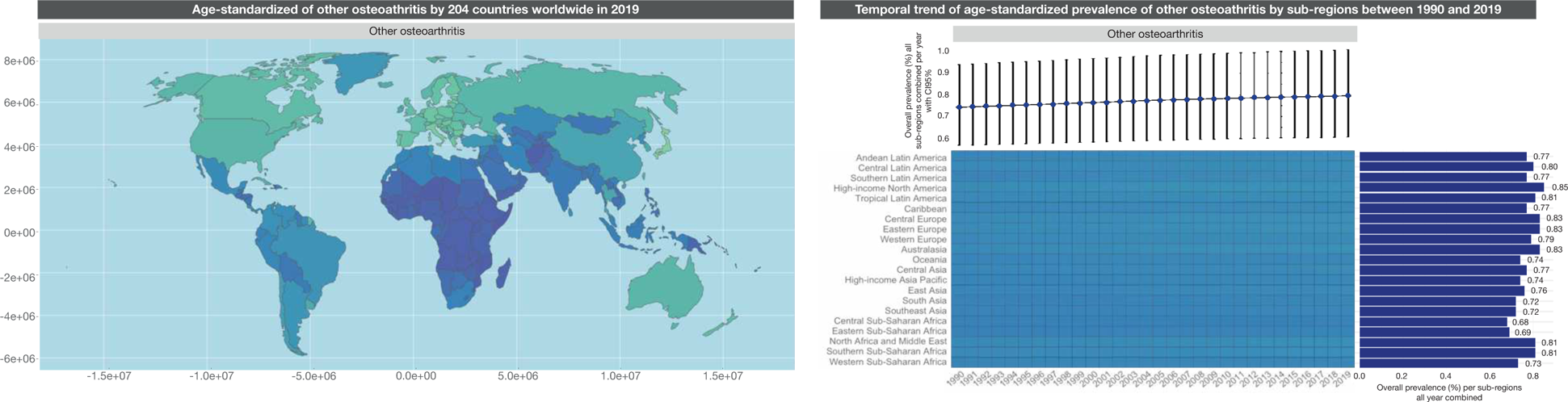
Spatial and temporal distribution of the age-standardized prevalence of MSD worldwide by 204 countries and 21 sub-regions between 1990 and 2019.

**S2:** The global prevalence of MSD considering their 95% CIs worldwide for the year 2019

| MSD | Prevalence (%) | CI 95% |
| --- | --- | --- |
| Low back pain | 8.62 | [7.62-9.74] |
| Neck pain | 2.45 | [1.95-3.10] |
| Osteoarthritis hip | 0.56 | [0.43-0.70] |
| Osteoarthritis knee | 5.02 | [4.32-5.77] |
| Osteoarthritis hand | 2.61 | [1.96-3.44] |
| Other osteoarthritis | 0.70 | [0.53-0.89] |

S3: The minimum and maximum prevalence of MSD considering their 95% CIs per country for the year 2019

**S3:** The minimum and maximum prevalence of MSD considering their 95% CIs per country for the year 2019

| Min/<br>max | MSD | Raw prevalence of MSD |  |  | Age-standardized prevalence of MSD |  |  |
| --- | --- | --- | --- | --- | --- | --- | --- |
|  |  | Country | Prevalence (%) | CI 95% | Country | Prevalence (%) | CI 95% |
| Minimum | Low back pain | India | 3.31 | [2.89-3.77] | India | 5.37 | [4.73-6.09] |
|  | Neck pain | Niger | 0.56 | [0.44-0.73] | New Zealand | 0.96 | [0.77-1.20] |
|  | Osteoarthritis hip | Afghanistan | 0.07 | [0.06-0.09] | Democratic People's Republic of Korea | 0.19 | [0.14-0.24] |
|  | Osteoarthritis knee | Somalia | 0.98 | [0.82-1.14] | Tajikistan | 2.77 | [2.38-3.21] |
|  | Osteoarthritis hand | Timor-Leste | 0.04 | [0.02-0.05] | Maldives | 0.12 | [0.08-0.16] |
|  | Other osteoarthritis | Somalia | 0.21 | [0.16-0.28] | Central African Republic | 0.67 | [0.51-0.86] |
|  | Average minimum prevalence |  | 0.86 | [0.73-1.01] | Average minimum prevalence | 1.68 | [1.43-1.96] |
| Maximum | Low back pain | Japan | 18.9 | [16.7-21.4] | United States of America | 13.7 | [12.7-14.9] |
|  | Neck pain | United States of America | 7.98 | [6.63-9.56] | Philippines | 5.55 | [4.42-7.01] |
|  | Osteoarthritis hip | Monaco | 2.08 | [1.57-2.64] | United States of America | 1.12 | [0.89-1.37] |
|  | Osteoarthritis knee | Japan | 16.9 | [19.1-14.7] | Republic of Korea | 6.74 | [5.80-7.71] |
|  | Osteoarthritis hand | Republic of Korea | 13.3 | [10.0-17.3] | Iceland | 6.21 | [4.65-8.06] |
|  | Other osteoarthritis | Monaco | 1.82 | [1.41-2.29] | Qatar | 1.01 | [0.75-1.30] |
|  | Average maximum prevalence |  | 10.16 | [10.5-11.3] | Average maximum prevalence | 5.72 | [4.65-6.72] |

**S4:**
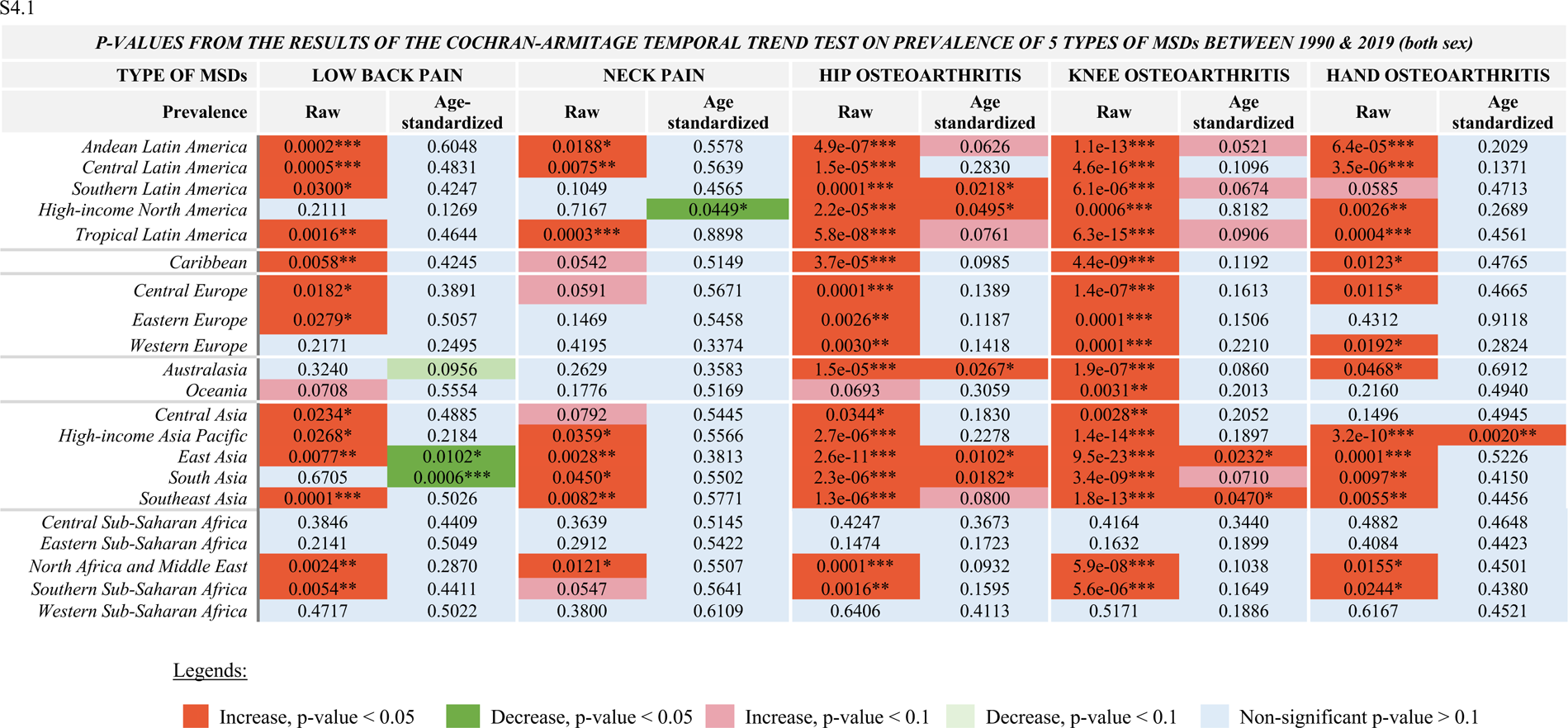
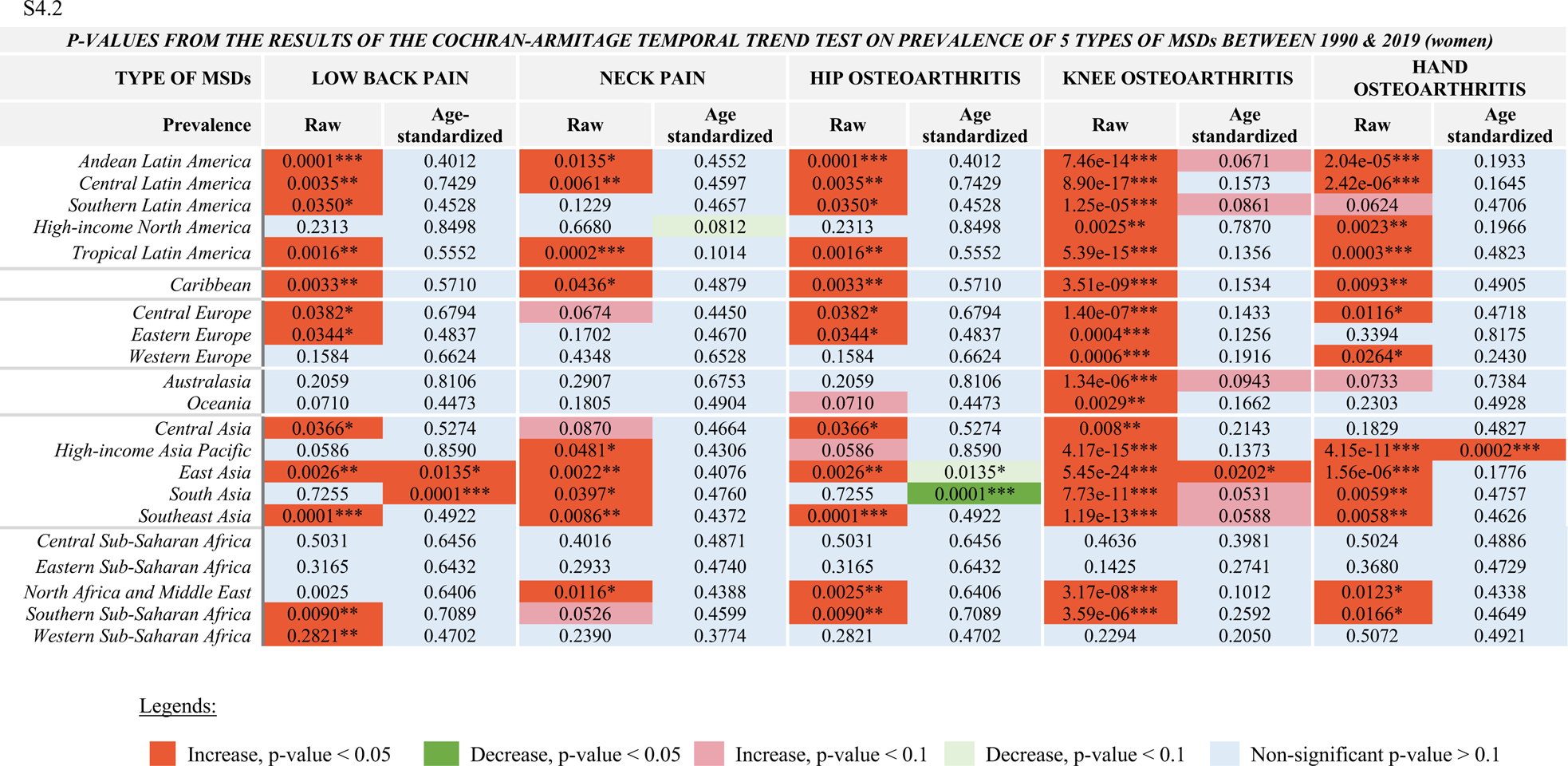
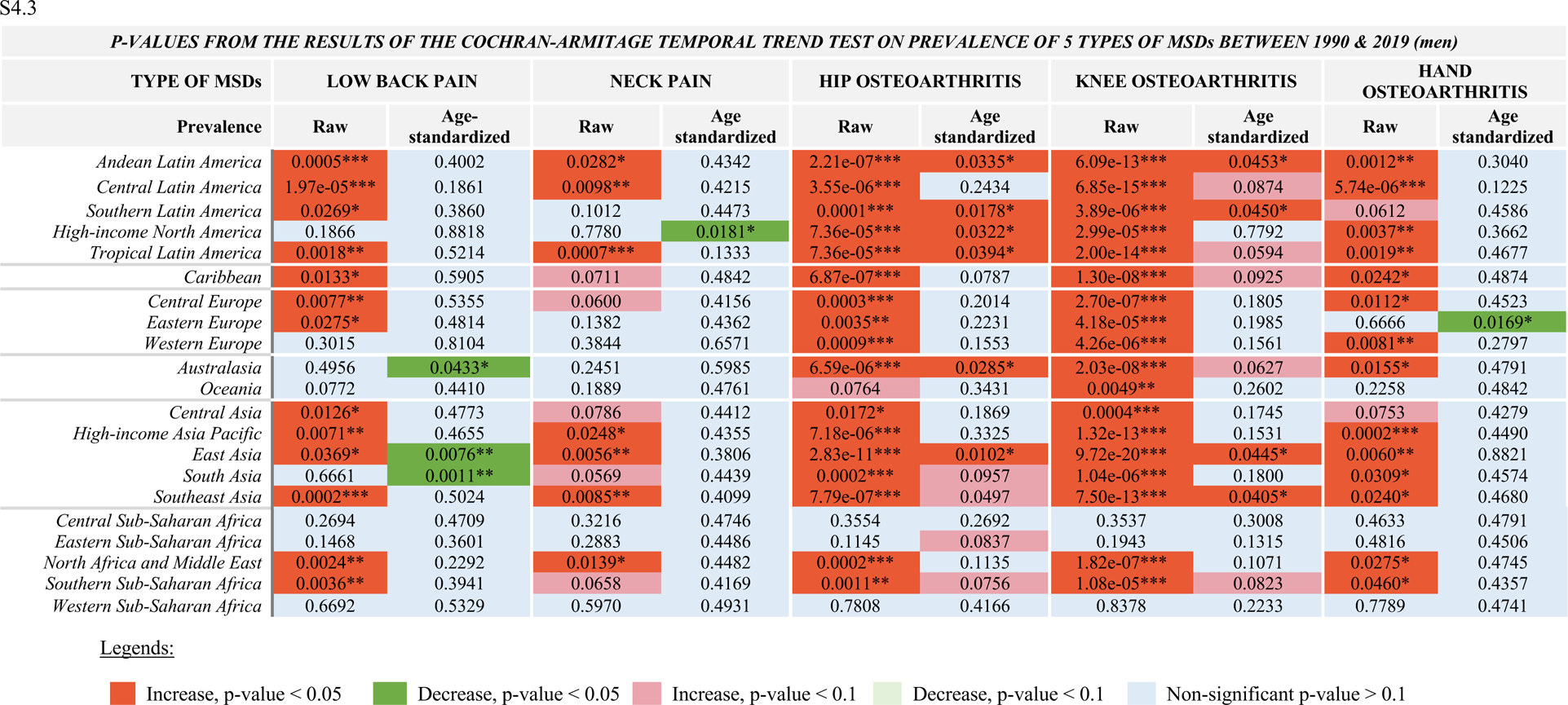
Details of the p-values from the Cochran-Armitage temporal trend tests on the prevalence of MSD between 1990 and 2019: in both sex (S4.1), women (S4.2), and men (S4.3). S4.1

**S5:** Linear regression models on 5 MSD for 2019. The World Bank income was an ordinal variable classified into four income levels: World Bank high-income, World Bank upper-middle-income, World Bank lower-middle-income, and World Bank low-income. The medical density variable for 1,000 individuals per country in 2017 was initially continuous and then categorized as an ordinal variable according to the highlighted four quantiles (Q1, Q2, Q3 and Q4).

|  | Low back pain |  |  | Neck pain |  |  | Hip osteoarthritis |  |  | Knee osteoarthritis |  |  | Hand osteoarthritis |  |  |
| --- | --- | --- | --- | --- | --- | --- | --- | --- | --- | --- | --- | --- | --- | --- | --- |
| Predictors | Estimates | CI (95%) | P-Value | Estimates | CI (95%) | P-Value | Estimates | CI (95%) | P-Value | Estimates | CI (95%) | P-Value | Estimates | CI (95%) | P-Value |
| World Bank Incomes | 1.54 | [1.16 – 1.92] | <0.001 | 0.55 | [0.31 – 0.78] | <0.001 | 0.17 | [0.12 – 0.21] | <0.001 | 1.15 | [0.89 – 1.42] | <0.001 | 0.60 | [0.29 – 0.90] | <0.001 |
| Medical density | 0.71 | [0.51 – 0.92] | <0.001 | 0.20 | [0.07 – 0.33] | 0.002 | 0.12 | [0.10 – 0.15] | <0.001 | 0.55 | [0.41 – 0.69] | <0.001 | 0.52 | [0.35 – 0.69] | <0.001 |
| Observations | 187 |  |  | 187 |  |  | 187 |  |  | 187 |  |  | 187 |  |  |
| R<sup>2</sup>/R<sup>2</sup> adjusted | 0.654/0.650 |  |  | 0.340/0.333 |  |  | 0.688/0.685 |  |  | 0.697/0.694 |  |  | 0.469/0.463 |  |  |

